# Plasma vs. serum: which is better for proteomic blood biomarker analysis? Evaluation of the novel NULISA platform

**DOI:** 10.1101/2025.11.08.25339626

**Authors:** Marissa F. Farinas, Yijun Chen, Xuemei Zeng, Michel N. Nafash, Ann D. Cohen, Oscar I. Lopez, Thomas K. Karikari

## Abstract

**INTRODUCTION:** The NULISASeq™ CNS Disease Panel has high potential for AD diagnosis, but the comparability of serum vs. plasma remains unclear.

**METHODS:** We compared its performance on 43 matched serum-plasma pairs from a memory clinic cohort.

**RESULTS:** The panel reproducibly quantified 124 targets (mean CV=4.9%) with high detectability (mean=95.7%). Serum-plasma correlations were strong (ρ>0.7) for 79 targets. 48 targets had significant NPQ differences, with 32 higher in plasma. Plasma had more erythrocyte-enriched proteins (HBA1, PGK1, SOD1, PRDX6), while serum had more platelet-derived proteins (CD40LG, BDNF, VEGFA, Aβ40). For classical AD biomarkers, serum-plasma correlations were stronger for p-tau, GFAP, and NfL (ρ>0.9) than for Aβ targets (ρ=0.594– 0.785). Tau levels were higher in plasma; GFAP and NfL were similar, and Aβ peptides were mixed. Twelve targets, along with p-tau217/Aβ42, were linked to AD diagnosis, with plasma generally showing stronger effects.

**DISCUSSION:** Our results support serum use but suggest plasma performs better for AD using this panel.

## 1. BACKGROUND

The blood proteome, comprising proteins secreted from various tissues, holds promise for real-time monitoring of (patho)physiological states [1, 2]. With the advent of ultrasensitive immunoassays and mass spectrometry-based techniques, it is now possible to detect brain pathophysiology, including Alzheimer’s disease (AD), through peripheral blood biomarkers, even when they are present at very low concentrations [3, 4]. For example, several blood biomarkers, such as phosphorylated tau species, brain-derived tau (BD-tau), amyloid beta (Aβ) peptides, neurofilament light chain (NfL), and glial fibrillary acidic protein (GFAP), have shown strong potential for AD diagnosis [5–7]. Notably, Lumipulse p-tau217/Aβ42 ratio has recently been FDA-cleared for clinical diagnosis of Aβ pathology [8].

Serum and plasma, both representing the liquid components of blood, are widely used for biomarker measurements. However, they differ in composition due to distinct processing methods [9]. Plasma is obtained by centrifuging blood treated with anticoagulants to prevent clotting and retains clotting factors. Serum, on the other hand, is collected after blood has clotted, resulting in the loss of these factors. Since plasma collection does not require the clotting process—which can delay sample processing and lead to the loss of proteins trapped in clots—it generally exhibits lower variability and reduced protein degradation or cellular release compared to serum [10–12]. Serum also tends to have more platelet-derived proteins due to platelet aggregation and release of their contents [13]. Therefore, plasma is considered more suitable for proteomic studies and was recommended by the Human Proteome Organization’s Plasma Proteome Project (HPPP) as the preferred sample type for such research [14]. On the other hand, due to historical factors and the ease of preparation and handling, more clinical assays have been developed using serum, making it the more commonly utilized specimen in clinical practice [15, 16].

The impact of matrix types on the blood proteome has been documented by numerous studies [17–19], including several studies evaluating this effect specifically in the context of AD biomarkers, with most focusing on single-plex or low-plex immunoassays [6, 20–23]. While AD biomarker levels in serum and plasma are generally well correlated, certain biomarkers— particularly various tau and p-tau forms and Aβ peptides—consistently appear at lower levels in serum [6, 20–23]. This discrepancy is likely due to protein loss during the clotting process, which can trap or degrade analytes. Despite comparable diagnostic performance between serum and plasma for these biomarkers [24], the reduced concentrations observed in serum may limit its utility, especially in clinical research focused on preclinical stages of AD.

NUcleic acid Linked Immuno-Sandwich Assay (NULISA) is a recently developed ultrasensitive multiplex immunoassay that enables the simultaneous detection of many low-abundance proteins [25]. One of its multiplex panels, the NULISAseq™ CNS Disease Panel 120 (hereafter referred to as the NULISAseq CNS panel), enables the detection of over 120 key proteins implicated in AD pathogenesis. These include biomarkers associated with core amyloid/tau/neurodegeneration (AT[N]) pathology, as well as markers of mixed pathologies such as inflammation (I), vascular dysfunction (V), and synucleinopathy (S). Although it is still a relatively new technology, biomarkers measured using this NULISAseq CNS panel have demonstrated robust performance in detecting abnormal brain physiology associated with AD [26–29].

Given its high sensitivity, low sample volume requirements, and great clinical potential, the NULISAseq CNS panel is anticipated to become increasingly utilized in neurodegenerative disease biomarker research. Therefore, it is of critical importance to evaluate the impact of matrix type on the use of this panel. Toward this aim, in this study, we compared measurements and classification accuracies of NULISAseq CNS panel on paired plasma and serum from older adults (n=43) enrolled in an academic memory clinic.

## 2. METHODS

### 2.1 Cohort characteristics

We included 43 participants from the Alzheimer’s Disease Research Center (ADRC) at the University of Pittsburgh (Pitt-ADRC). This cohort takes part in ongoing annual clinical evaluations aimed at monitoring brain health over time and identifying potential disease biomarker changes. Participants were selected based on their sequence of clinical attendance and their informed consent, including authorization to collect blood samples specifically for this study. Each participant underwent an extensive neuropsychiatric evaluation that included medical history and physical/neurological examination, semi-structured psychiatric interview, neuroimaging, and neuropsychological assessments. All participants had adequate visual and auditory acuity to complete neuropsychological testing, and a reliable caregiver capable of providing correct information about clinical and cognitive symptoms. At the conclusion of these examinations, each individual set of results was reviewed by the study team (neurologists, neuropsychologists, and psychiatrists) at a consensus conference and provided a clinical diagnosis. To be diagnosed with probable AD, the patients must have had a gradual cognitive decline without history or evidence of illness that could cause mental impairment other than AD [30]. The ADRC study was reviewed and approved by the University of Pittsburgh Institutional Review Board (MOD19110245-023).

### 2.2 Blood collection and processing

Blood samples were collected according to the procedures described in Zeng et al. [31]. Briefly, blood was collected via venipuncture by experienced nurses trained in Pitt-ADRC procedures between 9:00 am and 2:00 pm with the time of last meal recorded. Whole blood from each participant was collected into a 10 mL lavender top EDTA tube and a 10 mL red top serum tube. Following each blood draw, the EDTA tubes were promptly inverted 8 to 10 times and red top tubes 5 times, left at room temperature for 30 minutes and centrifuged at 2000 xg for 10 minutes at 4°C. The resulting plasma and serum samples were aliquoted into cryovials and frozen at -80°C until use.

### 2.3 NULISAseq assay procedures and data processing

Paired plasma and serum samples were thawed and centrifuged at 4,000 xg for 10 min to remove particulates. The supernatants were analyzed using the NULISAseq CNS disease panel 120 (v2) on an Alamar ARGO^TM^ HT system as previously described [25]. Briefly, samples were incubated with a cocktail of paired capture and detection antibodies and internal control mCherry protein. The capture antibodies are conjugated with partially double-stranded DNA containing a poly-A tail and a target-specific barcode while the detection antibodies are conjugated with another partially double-stranded DNA containing a biotin group and a matching target-specific barcode. The immunocomplexes underwent magnetic bead-based capture, followed by washing, release into a low-salt buffer, recapture with streptavidin-coated magnetic beads, and a second round of washing. DNA reporter molecules containing unique target and sample-specific barcodes were then generated by ligation and quantified using next generation sequencing (NGS).

A total of 124 biomarkers were included in the NULISAseq CNS panel. Protein levels for each target were quantified by first normalizing the raw counts. This was achieved by dividing the target count for each sample well (plus one) by the corresponding mCherry internal control count (plus one). The resulting values were then rescaled and log -transformed to generate NULISA Protein Quantification (NPQ) units, which served as surrogate measures of protein abundance. Fold changes between serum and plasma were calculated as 2 to the power of the difference in NPQ. Ratios between two targets, in particularly Aβ42/Aβ40 and p-tau217/Aβ42, were represented by their NPQ value difference. A sample control was measured in triplicate to assess the reproducibility of the assay.

### 2.4 Statistical analysis

Analyses were performed using the R statistical software (version 4.2.1, R Foundation for Statistical Computing, Vienna, Austria) and MATLAB (R2024a, MathWorks), and plots were generated with R, MATLAB, or GraphPad Prism (version 10.2.3). Demographic data were summarized using medians and interquartile ranges (IQRs) for continuous variables, and frequencies with corresponding percentages for categorical variables. Spearman correlations measured the strength of associations of NULISAseq CNS panel biomarker levels between plasma and serum. Paired t-test was used to assess the significance of the differences in detectability and absolute NPQ values between plasma and serum.

Logistic regression was used to evaluate the predictive performance of NULISAseq CNS panel biomarkers for AD diagnosis. The clinical diagnosis of AD served as the dependent variable, while biomarker NPQ values were included as independent variables, adjusted for age and sex. The resulting models were used to calculate the area under the curve (AUC). The Delong test was used to compare AUC differences between plasma and serum.

The Benjamini–Hochberg Procedure was applied to adjust for multiple comparisons [32]. An adjusted p-value of less than 0.05 was considered statistically significant across all analyses.

## 3. RESULTS

### 3.1 Participant characteristics

This study included 43 matched plasma and serum samples collected simultaneously from Pitt-ADRC participants during their annual visits. Table 1 summarizes the demographic characteristics of the participants. The median age was 76.0 (70.0-81.5) years, and 18 (41.9%) were female. The majority of the cohort (81.4%) identified as non-Hispanic White. 14 participants (32.6%) were diagnosed with probable AD and 29 (67.4%) as normal controls. According to Clinical Dementia Rating global score (CDR-GS), 15 participants (34.9%) had no dementia (CDR-GS = 0), 19 (44.2%) had very mild dementia (CDR-GS = 0.5), 4 (9.3%) had mild dementia (CDR-GS = 1), and 5 (11.6%) had moderate dementia (CDR-GS = 2). Median Mini-Mental State Examination (MMSE) and Montreal Cognitive Assessment (MoCA) scores were 27.0 (23.0-29.0) and 25.0 (17.0-20.0), respectively. Probable AD was associated with higher CDR-GS (p = 0.001), and lower MMSE and MoCA scores, with both p < 0.001.

**Table 1.** Demographic characteristics of the study cohort.

|  | **Total** | **Normal control** | **Probable AD** | ***p-value*** |
| --- | --- | --- | --- | --- |
| **Overall** | 43 | 29 | 14 |  |
| **Age (years [IQR])** | 76.0 [70.0, 81.5] | 77.0 [70.0, 82.0] | 75.5 [70.3, 76.8] | 0.755 |
| **Sex (%)** |  |  |  |  |
| **Male** | 25 (58.1) | 17 (58.6) | 8 (57.1) | 1 |
| **Female** | 18 (41.9) | 12 (41.4) | 6 (42.9) |  |
| **Race (%)** |  |  |  |  |
| **Non-Hispanic White** | 35 (81.4) | 22 (75.9) | 13 (92.9) | 0.177 |
| **Black** | 5 (11.6) | 5 (17.2) | 0 (0.0) |  |
| **Asian** | 1 (2.3) | 0 (0.0) | 1 (7.1) |  |
| **American Indian or Alaskan native** | 1 (2.3) | 1 (3.4%) | 0 (0.0) |  |
| **NA** | 1 (2.3) | 1 (3.4%) | 0 (0.0) |  |
| **CDR-GS (%)** |  |  |  |  |
| **CDR-GS = 0** | 15 (34.9) | 15 (51.7) | 0 (0.0) | 0.001 |
| **CDR-GS = 0.5** | 19 (44.2) | 12 (41.4) | 7 (50.0) |  |
| **CDR-GS = 1** | 4 (9.3) | 0 (0.0) | 4 (28.6) |  |
| **CDR-GS = 2** | 5 (11.6) | 2 (6.9) | 3 (21.4) |  |
| **MMSE (IQR)** | 27.0 [23.0, 29.0] | 29.0 [27.0, 29.0] | 22.0 [19.3, 23.8] | <0.001 |
| **MoCA (IQR)** | 25.0 [17.0, 28.0] | 27.0 [25.0, 29.0] | 15.5 [14.0, 19.8] | <0.001 |

The Wilcoxon rank sum test was applied to numerical variables (age, MMSE, and MoCA) and Fisher's exact test to categorical variables (sex, race, and CDR Global Score) to evaluate statistical significance of the difference between the AD and normal control groups.
Abbreviation: NA, Not available; CDR-GS, Clinical Dementia Rating global score; MMSE, Mini-Mental State Examination; MoCA, Montreal Cognitive Assessment.

### 3.2 Detectability comparison between serum and plasma

A total of 124 protein targets were quantified using the NULISAseq CNS panel in this study. All 86 samples (43 plasma and 43 serum) were analyzed on a single plate. The assay demonstrated high reproducibility, with a median intra-plate coefficient of variation (CV) of 4.9%, based on triplicate measurements of the sample control (Figure 1A). Across combined serum and plasma samples, the assay showed a mean target detectability of 95.4% (±14.9%), defined as the percentage of samples exceeding the limit of detection (LOD) for each target, indicating high assay sensitivity. Of all targets included in the panel, only 23 showed detectability below 100% (Figure 1B), including three targets—pTDP43-409 (TDP43 phosphorylated at Ser409), SNCB (beta-synuclein), and PTN (pleiotrophin)—that showed detectability below 50%. Overall, plasma samples exhibited slightly but significantly higher target detectability compared to serum (p = 0.0037). 23 targets had detectability below 100% in serum, compared to 17 targets in plasma. Among the 23 targets with <100% detectability in combined samples, the majority (n=15) exhibited lower detectability in serum compared to plasma (Figure 1C).

**Figure 1.**
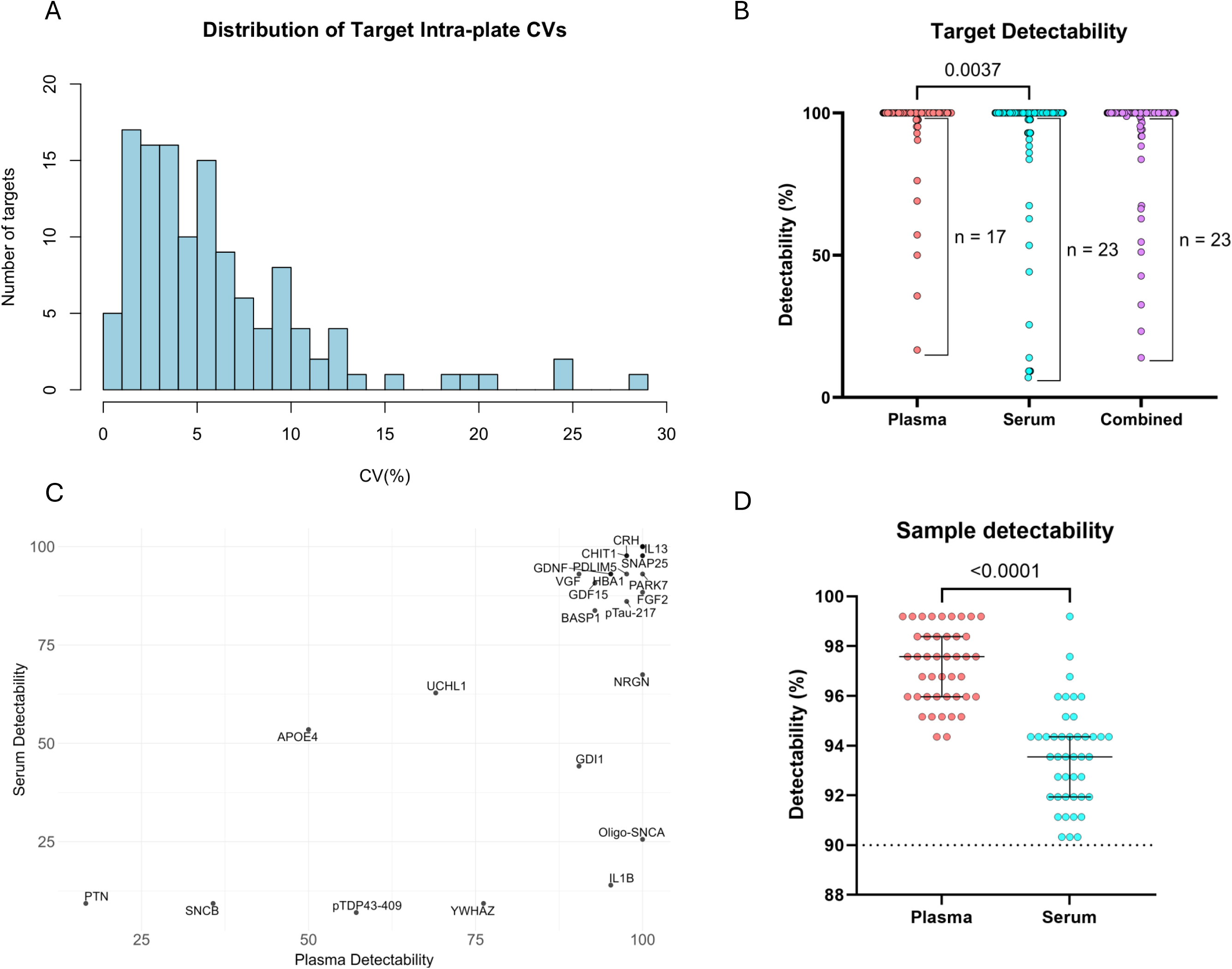
Analytical Performance of the NULISAseq CNS Panel in Serum and Plasma Samples. (A) Histogram showing intra-plate coefficient of variation (CV, %) for targets, based on triplicate analysis of a sample control. (B) Comparison of target detectability across sample types. (C) Scatterplot of target detectability in serum vs. plasma for targets with <100% detectability across all samples. (D) Comparison of sample detectability. Target detectability is defined as the number of samples in which a given target is above the limit of detection (LOD), while sample detectability refers to the number of targets above LOD within a given sample.

Similarly, we observed significantly higher overall sample detectability in plasma compared to serum, defined as the number of targets exceeding their respective limits of detection (LOD) per sample (p < 0.001; Figure 1D). Plasma samples had a mean sample detectability of 97.2% (±11.3%) compared to 93.6% (±20.1%) in serum.

### 3.3 Correlation between serum and plasma

We then evaluated the correlation of each target between plasma and serum using the Spearman correlation coefficient (ρ) to assess the strength of the association. As shown in Figure 2A, we observed an overall high correlation between the two matrix types, with a mean (±SD) ρ of 0.712 (±0.252). According to the cutoffs proposed by Schober et al. [32], 79 targets exhibited strong or very strong correlations (ρ ≥ 0.7), 25 showed moderate correlations (0.4 ≤ ρ ≤ 0.69), and 20 demonstrated either weak or no correlation (ρ < 0.4) (Figure 2A). Targets with the lowest correlation included NRGN (neurogranin; ρ = 0.076), YWHAZ (14-3-3 protein zeta/delta; ρ = 0.121), ENO (gamma-enolase; ρ = 0.132), IL7 (interleukin-7; ρ = 0.136), and UCHL1 (ubiquitin carboxy terminal hydrolase L1; ρ = 0.150).

**Figure 2.**
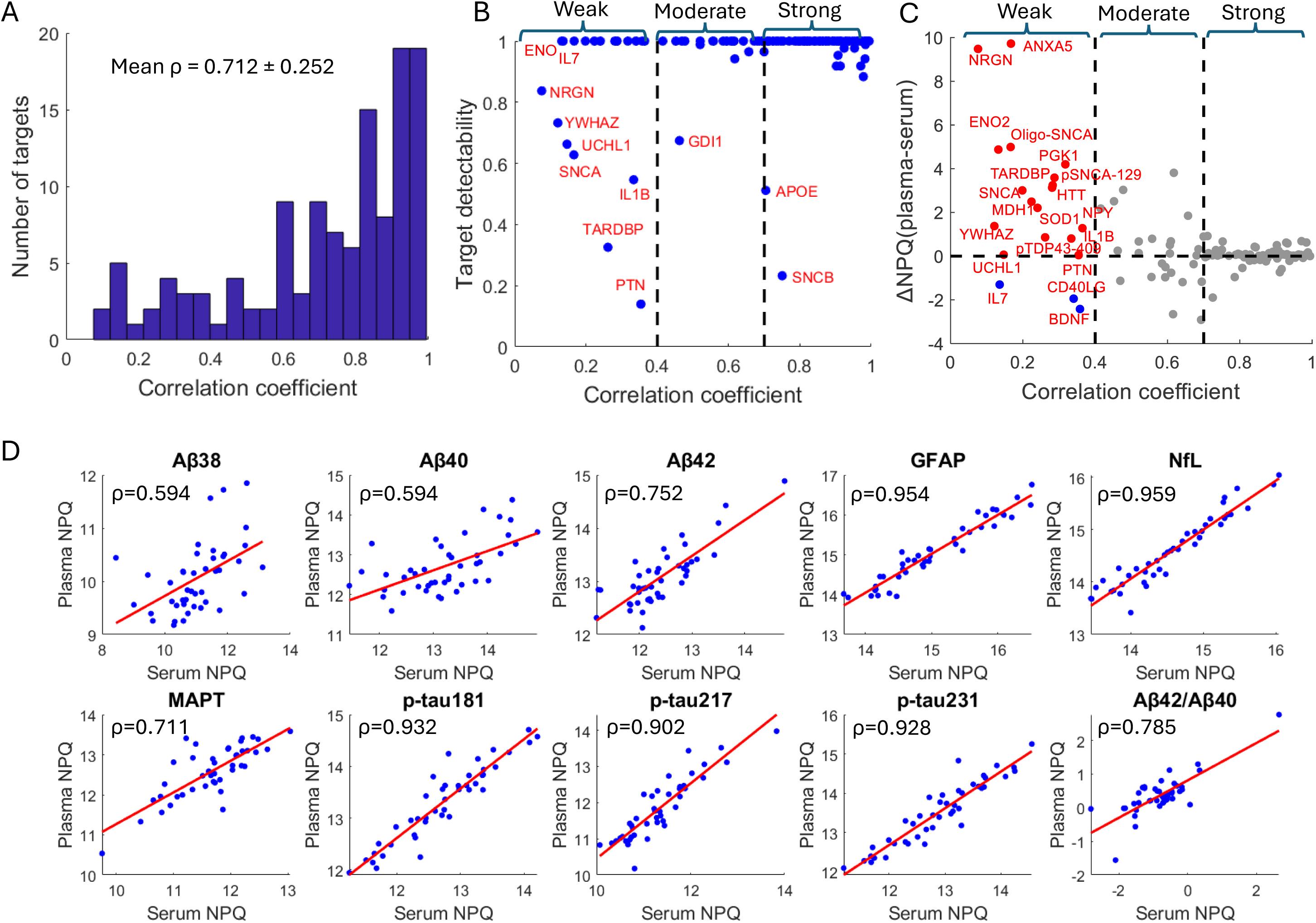
Correlation of NULISAseq CNS panel measurements between serum and plasma. (A) Histogram showing the distribution of Spearman correlation coefficients for all plasma–serum biomarker pairs. (B) Scatter plot illustrating the relationship between serum–plasma correlation (Spearman correlation coefficient; x-axis) and target detectability (y-axis). (C) Scatter plot illustrating the relationship between serum-plasma correlation and the mean NPQ difference between serum and plasma (ΔNPQps). (D) Correlation between serum and plasma levels for classical Alzheimer’s disease biomarkers. Red lines indicate linear regression lines.

We next investigated whether biomarkers with lower serum-plasma correlations were associated with other analytic metrics. As shown in Figure 2B, there was no significant agreement between correlation strength and target detectability (ρ = 0.123, p = 0.175). Notably, among the 20 targets exhibiting weak or no correlation, 13 showed 100% detectability in both serum and plasma. Interestingly, we observed a significant negative correlation between serum-plasma correlation and the mean NPQ difference (NPQ□□□□□□− NPQ□□□□□, denoted as ΔNPQ_ps_), with a ρ of −0.268 (p = 0.003). Out of the 20 targets with weak or no correlation, 17 had higher NPQ values in plasma.

We also evaluated the correlation between serum and plasma levels of classical AD biomarkers, including phosphorylated tau species, total tau (MAPT), Aβ peptides, neurofilament light chain (NfL), and glial fibrillary acidic protein (GFAP). As shown in Figure 2D, all phosphorylated tau species, as well as NfL and GFAP, exhibited very strong correlations between serum and plasma, with ρ values exceeding 0.9. Total tau (MAPT) also demonstrated a strong correlation, though slightly lower than p-tau species, with a ρ value of 0.711. In contrast, the serum-plasma correlation for Aβ peptides was generally weaker than for tau-related biomarkers. Among the Aβ peptides, only Aβ42 showed a strong correlation (ρ = 0.752), while Aβ38 and Aβ40 exhibited moderate correlations (both with a ρ value of 0.594). Aβ42/Aβ40 and p-tau217/Aβ42, two commonly used biomarker ratios for AD diagnosis, also showed strong correlations, with ρ values of 0.785 and 0.857, respectively.

### 3.4 Comparison of absolute levels of NULISAseq targets between plasma and serum

We utilized the paired t-test to evaluate the significance of differences in absolute NPQ values between serum and plasma. A total of 48 targets were differentially expressed (adjusted p < 0.05) and with mean NPQ value difference >0.5 (corresponding to NGS count difference of about 40%), with 32 elevated in plasma and 16 elevated in serum (Figure 3A and Table 2). As expected, there was a significant inverse association between serum-plasma correlation and the significance of NPQ value differences (ρ = -0.589; p < 0.001). However, some targets exhibited significant NPQ value differences despite strong serum-plasma correlations (Figure 3B). Among targets with significant NPQ value differences, 18, 24, and 32 of them showed weak, moderate, and strong serum-plasma correlations, respectively.

**Figure 3.**
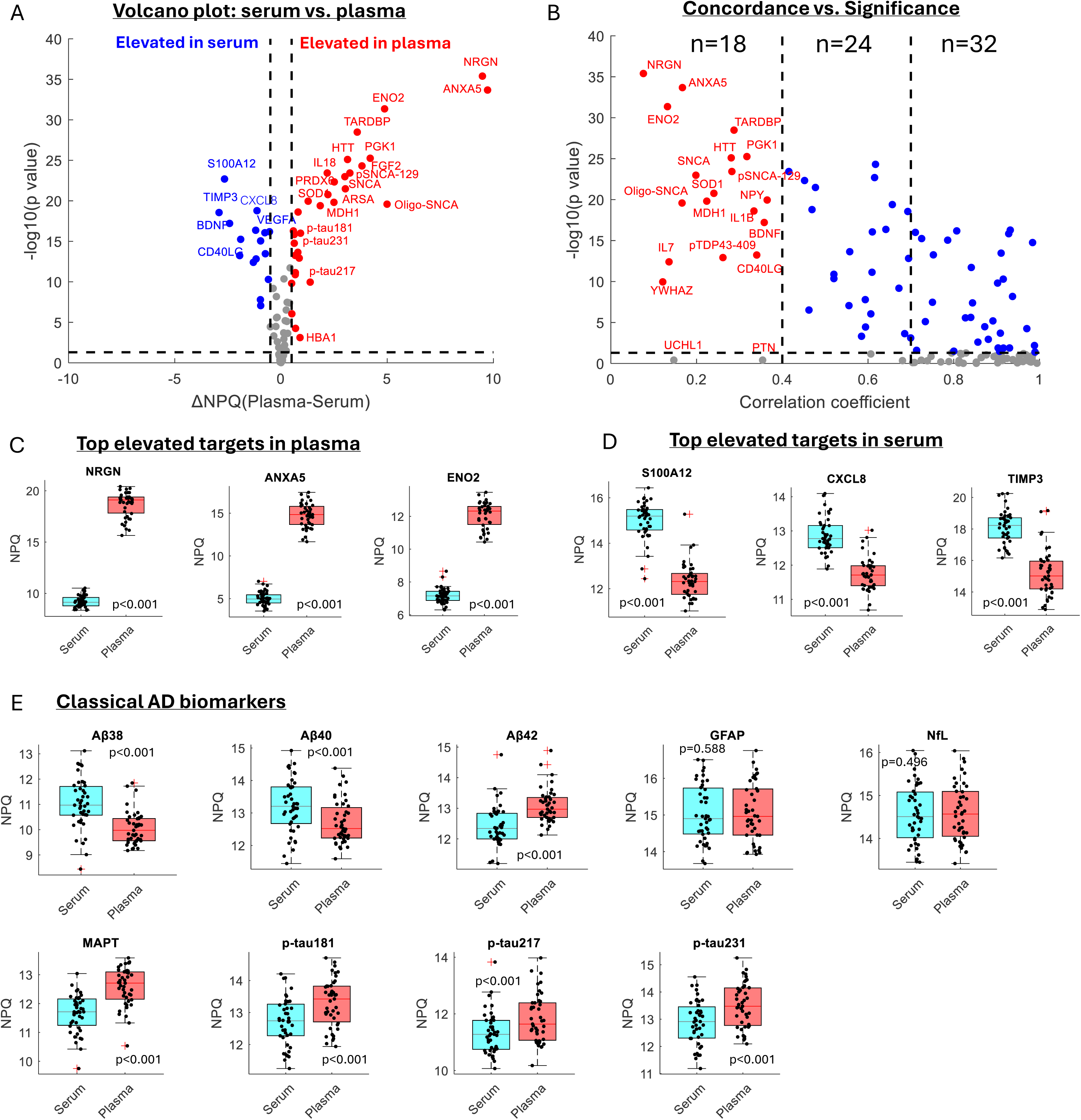
Differentially expressed proteins in plasma vs. serum. (A) Volcano plot showing the distribution of NPQ value differences and statistical significance. Red data points indicate targets significantly elevated in plasma, with a mean NPQ difference greater than 0.5. Blue data points represent targets significantly elevated in serum. Only selected significant targets were annotated with their target names. (B) Scatterplot illustrating the relationship between serum-plasma correlation (Spearman’s ρ) and the significance of NPQ value difference. Red datapoints represent targets with significant difference and weak serum-plasma correlation. Blue datapoints represent targets with significant correlation and moderate to strong serum-plasma correlation. (C) Boxplot distributions with overlaid individual data points for the top elevated targets in plasma. (D) Boxplot distributions with overlaid individual data points for the top elevated targets in serum. (E) Boxplot distributions with overlaid individual data points for classical AD biomarkers. P values are based on paired t-test, adjusted for multiple comparison using Benjamini–Hochberg Procedure.

**Table 2.**
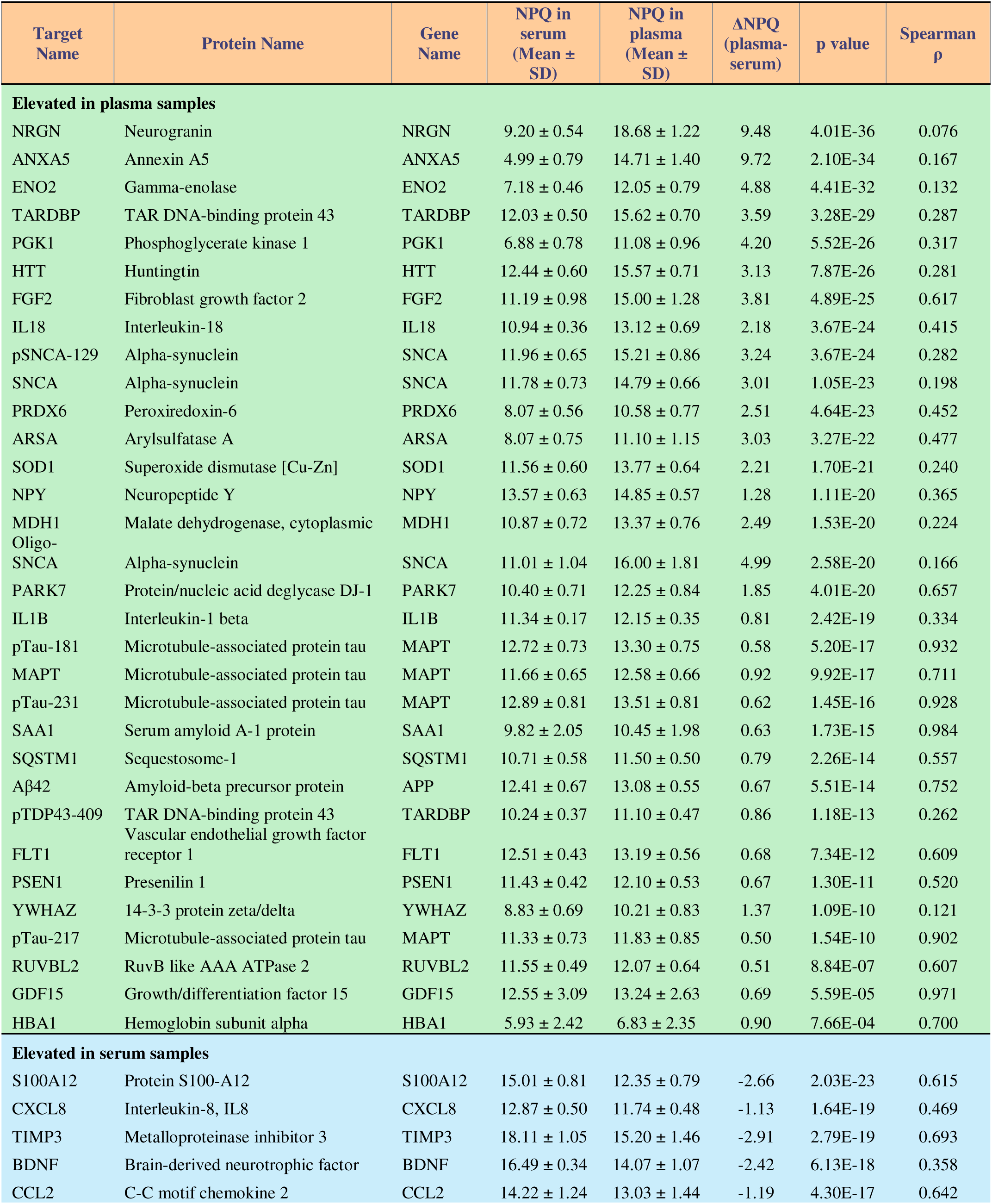
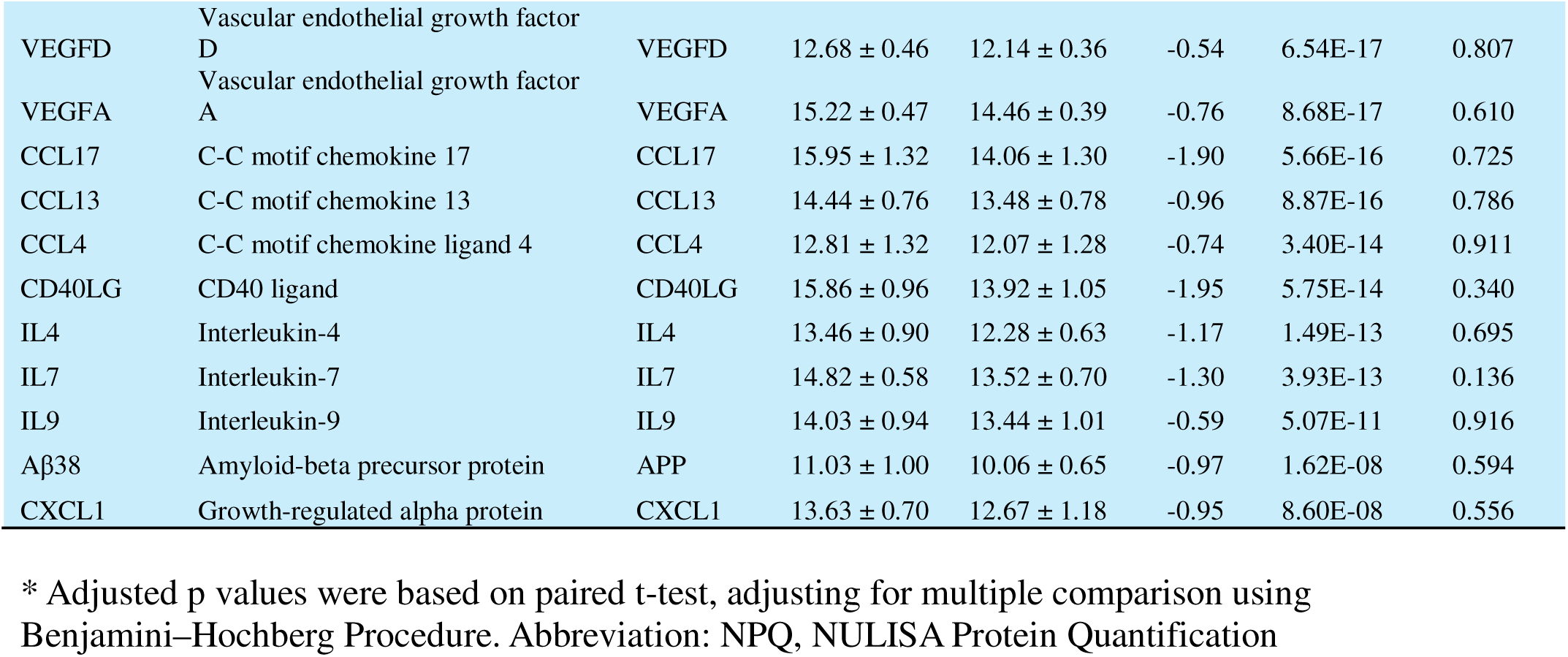
List of biomarkers with significant differences between serum and plasma.

Top significant targets elevated in plasma included NRGN (ΔNPQ_ps_ = 9.48), annexin A5 (ANXA5; ΔNPQ_ps_ = 9.72), ENO2 (ΔNPQ_ps_ = 4.88) and TAR DNA-binding protein 43 (TARDBP; ΔNPQ_ps_ = 3.59) (Figure 3C). In addition, all four proteins listed in the erythrocyte contamination panel by Geyer et al [34] including hemoglobin subunit alpha (HBA1), peroxiredoxin-6 (PRDX6), superoxide dismutase [Cu-Zn] (SOD1), and phosphoglycerate kinase 1 (PGK1), were significantly elevated in plasma, with ΔNPQps of 0.90, 2.51, 2.21, and 4.20, respectively (all with p < 0.001).

Protein S100-A12 (S100A12, Δ[plasma–serum] = -2.66), interleukin-8 (CXCL8; ΔNPQ_ps_ = -1.13), and metalloproteinase inhibitor 3 (TIMP3; ΔNPQ_ps_ = -2.91) were among the top significant targets elevated in serum (Figure 3D). All these targets had a p value < 0.0001.

Additionally, platelet-enriched targets, including CD40 ligand (CD40LG), brain-derived neurotrophic factor (BDNF), and vascular endothelial growth factor A (VEGFA), were found to be elevated in serum, with ΔNPQps of -1.95, -2.42, and -0.76, respectively (all with p < 0.001).

Figure 3E illustrates the NPQ value differences between serum and plasma for the classical AD biomarkers. All tau targets included in the NULISAseq CNS panel were significantly elevated in plasma, with ΔNPQ_ps_ of 0.92, 0.58, 0.50, and 0.62 for MAPT, p-tau181, p-tau217, and p-tau231, respectively (all p < 0.001). Interestingly, among the three Aβ peptides included in the panel, Aβ42 was significantly elevated in plasma (ΔNPQ_ps_ = 0.67, p < 0.001), whereas Aβ38 and Aβ40 were significantly decreased, with ΔNPQ_ps_ of -0.97 and -0.50, respectively, both with p value < 0.001. No significant differences were observed for GFAP and NfL levels between serum and plasma (p > 0.05).

### 3.5 Clinical performance of NULISAseq targets measured in plasma vs. serum

We then compared the clinical performance of NULISAseq targets in distinguishing participants clinically diagnosed with AD dementia from cognitively unimpaired (CU) individuals. As illustrated in Figures 4A and 4B, both serum and plasma samples revealed a similar set of targets with significant differences between the AD and CU groups, each identifying 11 targets with p values < 0.05, adjusted for multiple comparisons. Notably, Kallikrein-6 (KLK6) was uniquely significant in serum, while chemokine-like protein TAFA-5 (TAFA5) showed significance only in plasma samples. Table 3 summarizes the significant targets identified in both serum and plasma samples, along with their mean NPQ values by matrix type and their performance in distinguishing AD from CU. Among the two commonly used biomarker ratios, Aβ42/Aβ40 did not show significant associations in either serum or plasma. Notably, p-tau217/Aβ42 exhibited a significant association in plasma (p < 0.001), but not in serum (p = 0.07).

**Figure 4.**
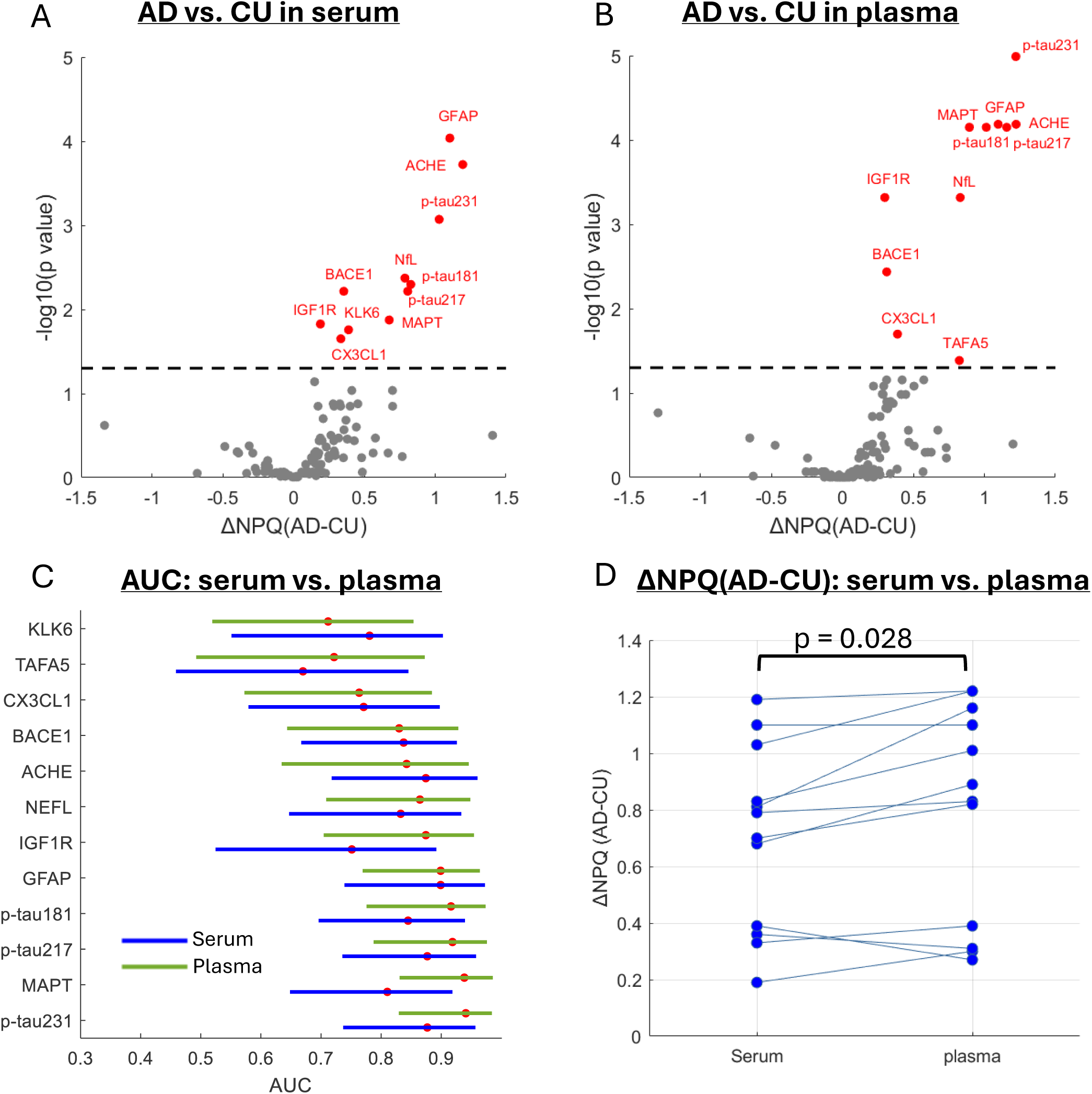
Clinical performance of NULISAseq CNS targets in distinguishing individuals with Alzheimer’s disease (AD) from cognitively unimpaired (CU) controls. (A, B) Volcano plots showing NPQ differences and statistical significance between AD and CU groups in serum (A) and plasma (B) samples. Red points indicate targets significantly elevated in AD (adjusted p < 0.05). P values were based on two-sided t test, adjusted for multiple comparisons. (C) Forest plot displaying AUC values and 95% confidence intervals for significant targets in plasma (green) and serum (blue). (D) Comparison of NPQ differences between AD and CU across serum and plasma for significant targets. Each blue dot represents an individual data point, with lines connecting corresponding serum and plasma values for the same target.

**Table 3.** List of targets significantly differentiating clinically diagnosed AD from cognitive unimpaired controls.

| Target | Serum |  |  |  | Plasma |  |  |  | AUC distinguishing AD and CU |  |  |
| --- | --- | --- | --- | --- | --- | --- | --- | --- | --- | --- | --- |
| | Mean (±SD) in AD | Mean (±SD) in CU | ΔNPQ<sub>ps</sub> | p value | Mean (±SD) in AD | Mean (±SD) in CU | ΔNPQ<sub>ps</sub> | p value | Serum | Plasma | DeLong p value |
| pTau-231 | 12.55±0.70 | 13.58±0.54 | 1.03 | 8.51E-04 | 13.11±0.63 | 14.33±0.44 | 1.22 | 1.02E-05 | 0.877 (0.737–0.957) | 0.941 (0.829–0.984) | 0.051 |
| ACHE | 12.99±0.51 | 14.19±0.94 | 1.19 | 1.89E-04 | 13.05±0.39 | 14.27±1.02 | 1.22 | 6.51E-05 | 0.874 (0.718–0.960) | 0.842 (0.634–0.946) | 0.376 |
| GFAP | 14.69±0.58 | 15.79±0.58 | 1.10 | 9.18E-05 | 14.71±0.62 | 15.81±0.56 | 1.10 | 6.51E-05 | 0.899 (0.739–0.973) | 0.899 (0.769–0.964) | 1.000 |
| pTau-217 | 11.07±0.69 | 11.87±0.50 | 0.81 | 6.09E-03 | 11.45±0.71 | 12.61±0.54 | 1.16 | 7.07E-05 | 0.877 (0.735–0.958) | 0.919 (0.787–0.976) | 0.161 |
| MAPT | 11.44±0.62 | 12.12±0.48 | 0.68 | 1.34E-02 | 12.29±0.59 | 13.19±0.27 | 0.89 | 7.07E-05 | 0.810 (0.648–0.919) | 0.938 (0.831–0.986) | 0.077 |
| pTau-181 | 12.45±0.69 | 13.28±0.47 | 0.83 | 5.04E-03 | 12.97±0.66 | 13.98±0.37 | 1.01 | 7.07E-05 | 0.845 (0.696–0.939) | 0.916 (0.776–0.974) | 0.046 |
| NEFL | 14.32±0.58 | 15.10±0.55 | 0.79 | 4.25E-03 | 14.33±0.56 | 15.16±0.51 | 0.83 | 4.82E-04 | 0.833 (0.647–0.933) | 0.865 (0.709–0.948) | 0.285 |
| IGF1R | 13.54±0.14 | 13.73±0.21 | 0.19 | 1.49E-02 | 13.49±0.13 | 13.79±0.29 | 0.30 | 4.82E-04 | 0.751 (0.525–0.892) | 0.874 (0.704–0.954) | 0.080 |
| BACE1 | 13.09±0.30 | 13.45±0.21 | 0.36 | 6.09E-03 | 13.12±0.26 | 13.43±0.19 | 0.31 | 3.66E-03 | 0.837 (0.667–0.926) | 0.830 (0.644–0.928) | 0.898 |
| CX3CL1 | 13.26±0.28 | 13.60±0.37 | 0.33 | 2.23E-02 | 13.26±0.30 | 13.65±0.44 | 0.39 | 2.00E-02 | 0.771 (0.579–0.897) | 0.764 (0.573–0.885) | 0.877 |
| TAF A5 | 13.20±0.56 | 13.89±1.14 | 0.70 | 9.17E-02 | 13.56±0.63 | 14.38±1.13 | 0.82 | 4.12E-02 | 0.670 (0.459–0.845) | 0.722 (0.492–0.873) | 0.321 |
| KLK6 | 12.67±0.31 | 13.06±0.43 | 0.39 | 1.75E-02 | 12.78±0.37 | 13.05±0.44 | 0.27 | 1.89E-01 | 0.781 (0.551–0.903) | 0.712 (0.519–0.854) | 0.148 |
| p-tau217/Aβ42 | (-1.28)±0.58 | (-0.67)±0.82 | 0.61 | 7.00E-02 | (-1.60)±0.62 | (-0.53)±0.85 | 1.07 | 4.64E-04 | 0.739 (0.541–0.876) | 0.50 (0.647–0.951) | 0.012 |

P values for NPQ value differences between AD and CU were calculated using a two-sided t-test and adjusted for multiple comparisons via the Benjamini–Hochberg procedure. AUC comparisons were performed using the DeLong test. The p values shown are unadjusted for multiple comparisons. P-tau217/Aβ42 was represented by NPQ value difference between p-tau217 and Aβ42. Abbreviation: AD, Alzheimer's disease; CU, cognitive unimpaired; SD, standard deviation; ΔNPQ<sub>AC</sub>, mean NPQ difference between AD and CU.

Top-performing targets included several classical AD biomarkers, such as total tau (MAPT), various p-tau forms, GFAP, and NfL. Among plasma samples, p-tau231 demonstrated the highest accuracy in distinguishing AD from CU, with an AUC of 0.941 (95% CI: 0.829– 0.984). This was closely followed by total tau, p-tau217, and p-tau181, with AUCs of 0.938 (95% CI: 0.831–0.986), 0.919 (95% CI: 0.787–0.976), and 0.916 (95% CI: 0.776–0.974), respectively. In serum samples, GFAP was the top-performing target (AUC = 0.899, 95% CI: 0.739–0.973), followed by p-tau231 and p-tau217, both with AUCs of 0.877. Top significant targets also included proteins with critical roles in AD pathology, such as acetylcholinesterase (ACHE) and beta-secretase 1, both elevated in AD, regardless of matrix type.

Overall, plasma samples yielded higher AUCs than their serum counterparts (Figure 4C). Among the biomarkers, p-tau181, p-tau231 and p-tau217/Aβ42 exhibited relatively larger differences between plasma and serum, with marginal statistical significance prior to correction for multiple comparisons (Table 3). However, after adjusting for multiple testing, none of the biomarkers showed statistically significant differences. Plasma samples also demonstrated greater effect sizes for these targets compared to serum, as illustrated in the scatterplot of NPQ differences between AD and CU (ΔNPQ_AC_) in Figure 4D. Of the 12 individual significant targets analyzed, 9 showed higher ΔNPQ_AC_ values in plasma than in serum. P-tau217/Aβ42 also showed a larger change between AD and CU in plasma than serum, with mean difference of 1.07 and 0.61, respectively (Table 3).

## 4. DISCUSSION

Historically, the development of blood-based biomarkers for AD has primarily focused on EDTA plasma, which has demonstrated robust clinical performance across multiple assay platforms [35–38]. In contrast, serum is the most commonly used matrix in clinical laboratories due to its ease of handling, absence of anticoagulants—which can interfere with routine blood tests and immunoassays—and the availability of long-established reference standards [16, 39, 40]. The NULISAseq CNS Disease Panel is a recent innovation in the AD blood biomarker field, integrating immunoassay-based detection with next-generation sequencing to achieve enhanced sensitivity and multiplexing capabilities, enabling the detection of over 120 key proteins involved in neurodegeneration. However, to the authors’ knowledge, no studies have yet investigated the potential impact of matrix type on the performance of this platform. The present study aims to address this gap.

Our findings demonstrate that the NULISAseq CNS panel achieves high detectability in both serum and plasma samples. The mean detectability exceeded 95%, with only three proteins—pTDP43-409, SNCB, and PTN—showing detectability below 50%. These results align with previous NULISA studies [27, 31, 41]. Furthermore, we observed strong overall correlations between serum and plasma, with the majority of targets (104 out of 124) exhibiting moderate to very strong correlations (ρ ≥ 0.4). Classical Alzheimer’s disease biomarkers— including various tau isoforms, GFAP, NfL, and Aβ peptides—also showed moderate to strong inter-matrix correlations. Notably, both serum and plasma samples revealed a similar subset of targets with significantly different abundance between individuals with clinical AD and cognitively unimpaired controls. Of the 12 combined significant targets, only two (TAFA5 and KLK6) were uniquely significant in one matrix type, based on an adjusted p value threshold of < 0.05. These findings support the panel’s robustness and suitability for blood-based AD biomarker applications across both matrices.

Despite the overall comparable performance between matrices, our results support plasma as the preferred matrix for the NULISAseq CNS panel. A substantial number of targets (n = 48) showed statistically significant and relatively large differences in absolute NPQ values, defined by an arbitrary threshold of 0.5 which corresponds to approximately a 40% difference in protein abundance after back-transforming the log2-transformed NPQ values. Notably, two-thirds of these biomarkers had higher NPQ values in plasma samples. Plasma also demonstrated higher classification accuracy in distinguishing AD from cognitively unimpaired individuals, although the difference was not statistically significant compared to serum based on the DeLong test. Additionally, plasma samples demonstrated greater effect sizes, as indicated by larger NPQ differences between the AD and CU groups compared to serum. Noticeably, the p-tau217/Aβ42 ratio, whose counterpart on the Lumipulse platform is FDA-approved for clinical use, only showed significance in plasma but not serum.

The primary distinction between serum and plasma processing lies in the clotting process, which contributes to differences in blood cell contamination, platelet protein release, and protein loss due to clot trapping. Our findings align with the expected lower blood cell contamination in serum, as erythrocytes become trapped within the clot. All four erythrocyte contaminant signature proteins [34], HBA1, PRDX6, SOD1, and PGK1, were significantly elevated in plasma. Our results also support higher levels of platelet protein contamination in serum, attributable to platelet activation and subsequent protein release during clot formation. Consistent with this, we observed several platelet-enriched targets, such as CD40LG [42], BDNF [43], and VEGFA [44, 45], being elevated in serum compared to plasma.

Among classical AD biomarkers, all tau targets were significantly elevated in plasma, consistent with findings from several previous comparative studies using alternative platforms [20, 21, 23]. Our results suggested comparable GFAP and NfL levels between serum and plasma. Prior studies on NfL have reported mixed results, with some indicating higher levels in serum, others in plasma, and some showing no significant difference between the two matrices [20, 23, 46]. To our knowledge, there are limited studies that examine the difference between serum and plasma GFAP levels. Our observation for equivalent level of GFAP agreed with Ashton et al. [23] who also observed similar GFAP in serum and plasma. Huebschmann et al. [47] reported elevated levels of plasma GFAP compared to serum GFAP. However, the study utilized different assays, although both with the Quanterix Simoa platform, and at different times. These discrepancies highlight the need for further investigation to clarify matrix-specific differences in GFAP and NfL quantification.

Unlike tau targets, we observed different trend for Aβ targets, with Aβ42 being elevated in plasma, whereas Aβ40 (ΔNPQps = -0.499, p < 0.001; not listed in Table 2 due to being just below the -0.5 cutoff) elevated in serum. We hypothesize that this differential pattern may be influenced by platelet activation and bursting during the clotting process. Platelets, contributing to over 90% of peripheral Aβ peptides, may release APP fragments or Aβ peptides during clotting [48]. Given that Aβ40 is the predominant isoform released by platelets, it may be more strongly affected by this platelet activation burst.

During clot formation, proteins can become sequestered within the clot matrix, leading to reduced serum levels [49]. We observed a greater number of proteins with significant NPQ differences elevated in plasma samples, suggesting that protein trapping may be a predominant factor contributing to serum–plasma proteomic differences. This loss of proteins during clotting likely increases variability in serum measurements, thereby diminishing serum–plasma correlation. It may also explain the observation that targets with weak serum–plasma correlations tend to exhibit higher levels in plasma.

This study has several notable strengths, including the use of a well-defined clinical cohort, the availability of matched plasma and serum samples, and the implementation of a multiplexed proteomic platform characterized by a wide dynamic range and high sensitivity. Nonetheless, certain limitations should be considered. First, the relatively small sample size (n = 43) reduces statistical power for subgroup analyses and may limit the robustness and reproducibility of the diagnostic model findings. Second, cross-sectional design limits the ability to assess longitudinal changes in biomarkers or evaluate their prognostic value. Third, our cohort lacked racial and ethnic diversity, and findings may not generalize to other populations.

In conclusion, our results demonstrate that serum performs comparably to plasma for many neurodegenerative disease biomarkers measured using the NULISAseq platform. While plasma retains marginal advantages in detectability and diagnostic performance for a subset of targets, serum remains a clinically viable matrix, particularly given its accessibility and routine use in clinical laboratories. Future studies with larger, longitudinal, and more diverse cohorts are warranted to confirm these findings, evaluate temporal dynamics, and further explore novel biomarker candidates that may be matrix specific.

## Data Availability

All data produced in the present study are available upon reasonable request to the authors from qualified investigators for the purpose of replicating the results in this manuscript, provided all applicable legal, ethical and institutional regulations are followed.

## ACKNOWLEDGEMENTS

We thank the Pitt-ADRC participants and their families and caregivers.

## CONSENT STATEMENT

All participants provided written consent, and the University of Pittsburgh Institutional Review Board approved the study.

